# Survival and outcomes of tocilizumab use in severe and critically ill COVID-19 patients not responding to steroids

**DOI:** 10.1101/2021.12.06.21266950

**Authors:** Shiraz Assu, Deepak Bhasin, Kavita Sekhri, Supriya Sampley, Harpal Singh, Gurleen Kaur

## Abstract

**Background:** Mortality and morbidity are highest in severe and critically ill patients with COVID -19 pneumonia. We used interleukin-6 (IL-6) receptor inhibitor, tocilizumab in the above patients who failed to show any clinical improvement after initial treatment with steroids.

**Methods:** This is a retrospective observational study conducted at a tertiary care hospital in India. Severe and critical COVID-19 patients, who got admitted to intensive care unit and subsequently received tocilizumab were included. Patients who worsened clinically or had no change in oxygen requirement even after 24hrs of receiving intravenous methylprednisolone at a dose of 1-2mg/kg/day received a maximum total dose of 800mg of intravenous tocilizumab. The day 28 all-cause mortality and progression to mechanical ventilation were the primary outcome measures. Clinical improvement and oxygen requirements after tocilizumab administration along with trends in inflammatory markers and secondary infections rates were also noted.

**Results:** A total of 51 patients who did not show clinical improvement even after 24 hours of intravenous steroids had received tocilizumab. In these patients, there was a significant decrease in oxygen requirement and clinical progression by day 7 of tocilizumab administration. Baseline median values of CRP (114.2 mg/L), IL-6 (69.8 pg/ml) and neutrophil-lymphocyte ratio (12.4) in these patients were elevated. Out of these, only CRP showed a significant decrease after the drug administration. 13 (26.5%) of the 49 patients on non-invasive oxygen support progressed to mechanical ventilation and the day 28 all-cause mortality was 10/51(19.6%). 10(19.6%) of the 51 patients had life threatening infections.

**Conclusion:** Early and timely administration of tocilizumab is a viable option in selected severe and critical COVID-19 patients who do not respond to initial steroids. When given along with steroids, a high suspicion of secondary infections should be kept.

## Introduction

A novel corona virus was first detected in Wuhan, in Hubei Province of China in late 2019.^1^ In February 2020, it was assigned the name COVID-19 by World Health Organization and on March 11th it was declared a pandemic. The corona virus study group of the International Committee on Taxonomy of Viruses (ICTV) has named it Severe Acute Respiratory Syndrome Corona virus 2 (SARS-COV-2).^2^ In spite of all the necessary measures taken to halt this emerging virus, it has spread worldwide.^3^ Due to pandemic and associated mortality, astronomical efforts are going on to discover molecules which effective against SARS-CoV-2. But even after more than one year of pandemic, wide scale research is still underway to reach a consensus regarding the perfect treatment protocol, which could significantly decrease the morbidity & mortality. One of the main reason behind this may be the unpredictability of the disease progression.

In view of the studied Covid immuno-pathogenesis, which showed the role of heightened cytokine release in COVID-19 associated systemic inflammation and hypoxic respiratory failure,^1, 4, 5^ many drugs altering these mechanisms have been tried as treatment. Among these, till date corticosteroids are the only medications that have shown to improve survival.^6-9^ This may be due to their broad spectrum anti-inflammatory effects. Another group which is being increasingly recognized are the immune modulators like monoclonal antibodies against interleukin-6 (IL-6) receptors, interleukin-1, CD6 etc. One of the most commonly studied and used drug among them is tocilizumab which inhibits the binding of IL-6 to both membrane and soluble IL-6 receptors. IL-6 is a pleotropic cytokine released in response to infection and stimulates inflammatory pathways as a part of the acute phase response. IL-6 inhibitors are currently being used as an anti-inflammatory agent in many conditions like rheumatoid arthritis, cytokine release syndrome due to chimeric antigen receptor T-cell therapy or giant cell arteritis. Infection by the severe acute respiratory syndrome-associated coronavirus (SARS-CoV) induces a dose-dependent production of IL-6 from bronchial epithelial cells.^10^

Few recent trials have shown good results with the use of tocilizumab in COVID-19. Based on these results, on June 26,2021 the United States Food and Drug Administration issued an emergency use authorization for tocilizumab, for the treatment of hospitalized adults band pediatric patients (>2 years) requiring supplemental oxygen. But in such patients, its timing of administration, dosage and effects are still unclear, especially when given in combination with corticosteroids.

To re-evaluate the role of tocilizumab when given along with steroids, we did this retrospective single center observational study on all severe and critical COVID-19 patients, who were administered tocilizumab.

## Materials and Methods

### Design and setting

We performed a retrospective study at a tertiary care hospital in Northern India. In view of the pure retrospective nature of the study, informed consent was waivered and the study was approved by the institutional scientific committee. (Ref no: TH/MSSH/MOHALI/RESEARCH)

Hospital electronic medical records were reviewed from April 2020 to February 2021. All consecutive adult (>18 years) patients confirmed as COVID-19 by reverse-transcription polymerase chain reaction assay or rapid antigen-positive test and admitted to the intensive care unit (ICU) with severe and critical disease [as defined by WHO(Table 1)], but without any evidence of sepsis and treated with tocilizumab were eligible for inclusion. The decision to administer tocilizumab was based on clinical worsening or lack of improvement in oxygen requirement even after >24hrs of receiving IV methylprednisolone at a dose of 1-2mg/kg/day, irrespective of the levels of various inflammatory markers. Patients were administered the drug only in the pulmonary or early hyper inflammatory phase (5-20 days from symptom onset) of the disease.^11^

**Table 1.** WHO severity definitions

|  |
| --- |
| <u>Critical COVID-19</u>: Defined by the criteria for acute respiratory distress syndrome (ARDS), sepsis, septic shock, or other conditions that would normally require the provision of life sustaining therapies such as mechanical ventilation (invasive or non-invasive) or vasopressor therapy. |
| <u>Severe COVID-19</u>: Defined by any of:<br>• Oxygen saturation < 90% on room air.<br>• Respiratory rate > 30 breaths/min.<br>• Signs of severe respiratory distress (accessory muscle use, inability to complete full sentences) |

Tocilizumab was not given to patients with known hypersensitivity to tocilizumab or any of their excipients, condition or treatment resulting in ongoing immunosuppression including neutropenia, known or suspected pregnancy, current evidence of any bacterial, fungal or other viral infections, previous history of tuberculosis, and patients with any of following laboratory results out of the ranges, as detailed below at screening.

a) Absolute neutrophil count ≤ 1.0 × 10^9^/L, b) Platelets < 50 × 10^9^/L, c) Aspartate aminotransferase or Alanine aminotransferase > 5x upper normal limit.

### Tocilizumab administration

Tocilizumab was given at a dose of 400mg intravenously as an infusion over 1 hour, followed by a second dose of up to 400mg (maximum total dose of 800mg) within 24 hours if there was no significant clinical improvement.

### Standard Treatment

Along with this all the patients also received standard treatment which included steroids (1-2mg/kg of methylprednisolone) for 5–7 days followed by tapering doses, antivirals (remdisivir 200mg on day 1, followed by 100mg once daily for 4 days), antibiotics at the discretion of the treating physician, prophylactic dose of low molecular weight heparin and other supportive management.

If not tolerating conventional oxygen support via nasal prongs or face mask, patients were given a trial of high flow nasal cannula (HFNC) and/or noninvasive ventilation (NIV) in ICU to achieve a SpO2 of 88–93%, if there was no contraindication. Patients were intubated and put on invasive ventilation, based on the following indications:

- Rapid progression of hypoxemia over hours.
- Unable to maintain oxygen saturation (SpO2) >88% on HFNC with a flow of 60 L/minute and FiO2 ≥90%
- Unable to maintain SpO2 > 88% on NIV with FiO2 ≥ 90% and/or persistent use of NIV for more ≥48 hours.
- Signs of respiratory fatigue (excessive use of accessory muscles of breathing), hypercarbia (pCO2 > 45 mm Hg), and/or altered mental status)
- Hemodynamic instability.
- Psychomotor agitation making nursing care impossible and requiring sedation.

### Outcome measures

The primary outcome were the incidence of progression to invasive mechanical ventilation and all-cause mortality at day 28.

The secondary outcome measures were clinical improvement [assessed using WHO ordinal scale for disease progression (Table 2)], change in oxygen requirements and inflammatory markers [C reactive protein (CRP), D-dimer, ferritin and neutrophil-lymphocyte ratio(NLR)] after tocilizumab administration. Factors predicting day 28 mortality, life threatening infections rates and other drug related side effects were also noted.

**Table 2.**
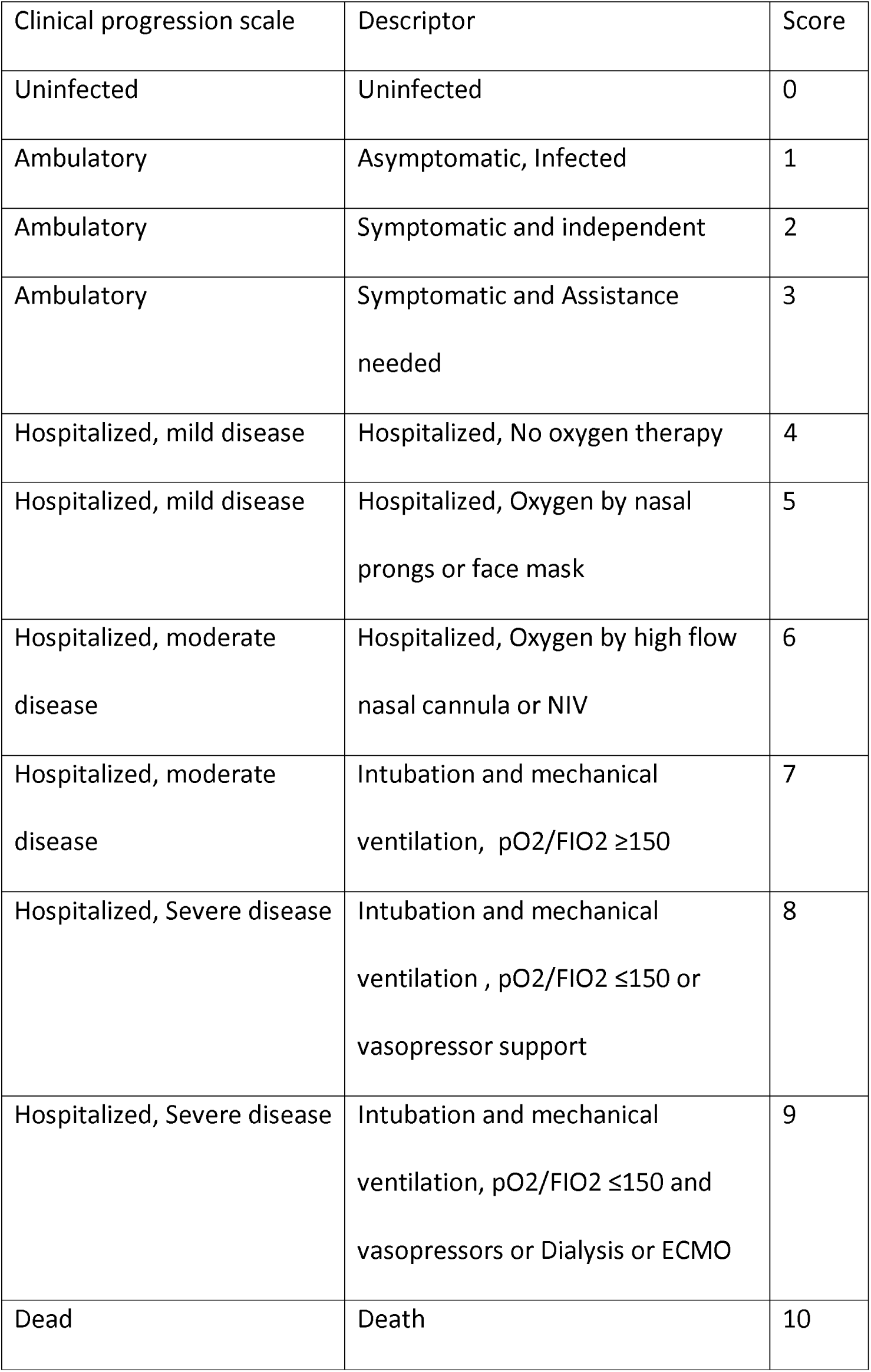
WHO clinical progression score(CPS)

### Statistical Analysis

All statistical analysis was done using SPSS software version 22. The descriptive statistics are reported as mean with standard deviation or median with interquartile ranges(IQR) for continuous variables and percentages/proportion for discrete variables. Wilcoxon–signed rank test was used to compare the non-normal quantitative values while Fisher’s exact test was applied for association between categorical variables. Logistic regression was used to find any independent factors associated with poor outcome after tocilizumab administration. All comparative tests were 2 tailed, and a p value of less than 0.05 was considered to be statistically significant.

## Results

A total of n=55 patients who did not show clinical improvement even after 24 hours of intravenous steroids were administered tocilizumab. Out of this 4 patients were excluded as they were shifted to other medical facility before day 28 or primary outcome. Hence further n=51 patients were included in the study. The mean age of the patients was 58[standard deviation(SD)-10.9] years, out of which 80% were males and 20% were females. The mean Body Mass Index (BMI) of the patients was 29kg/m^2^ (SD-5.7) with a majority of 41(80.4%) being overweight. Fever (86.3%) and cough (84.3%) were the most common symptoms, whereas hypertension (62.7%) followed by diabetes (58.8%) were the most common associated co-morbidities. The baseline characteristics of these patients are given in Table 3.

**Table 3.** Baseline demographics

|  |  |
| --- | --- |
| Age (years) |  |
| median(SD) | 58 (10.9) |
| Sex N(%) | Male – 41(80%)<br>Female – 10(20%) |
| BMI(kg/m2) |  |
| Mean (SD) | 29 (5.7) |
| Symptoms N(%) | Fever- 44(86.3%)<br>Cough – 43(84.3%)<br>Shortness of breath – 35(68.6%)<br>Myalgia – 9(17.6%)<br>Sore throat- 5(9.8%)<br>Diarrhea – 4(7.8%)<br>Vomiting- 3(5.9%)<br>Abdominal pain – 2(3.9%)<br>Oliguria -2(3.9%) |
| Co-morbidities N(%) |  |
| Hypertension | 32(62.7%) |
| Diabetes | 30(58.8%) |
| Coronary artery disease | 11(21.6%) |
| Chronic lung disease | 5(9.8%) |
| Chronic kidney disease | 4(7.8%) |
| Hypothyroid | 2(3.9%) |
| Others* | 8(15.7%) |

\*others – rheumatoid arthritis, parkinson’s disease, depression, pancreatitis, previous stroke, permanent pacemaker insitu; SD, standard deviation

Tocilizumab was administered at a median duration of 8 days since symptom onset and 2 days since hospitalization. At drug administration, 43 (84.3%) patients were receiving non-invasive respiratory support (Noninvasive ventilation or high flow nasal cannula), 6 (11.7%) patients were receiving conventional oxygen therapy (nasal prongs or venturi mask), and 2(3.9%) were on invasive mechanical ventilation. All of the patients (n=51) were administered tocilizumab based on clinical worsening >24 hours after steroid administration. More than one inflammatory marker was significantly raised in 40/51(78.4%) patients on the day of tocilizumab administration (Table 4). Out of the 46 patients in whom HRCT (high-resolution computed tomography) chest was available at tocilizumab administration, 32(69.5%) had CT severity index >15/25.

**Table 4.**
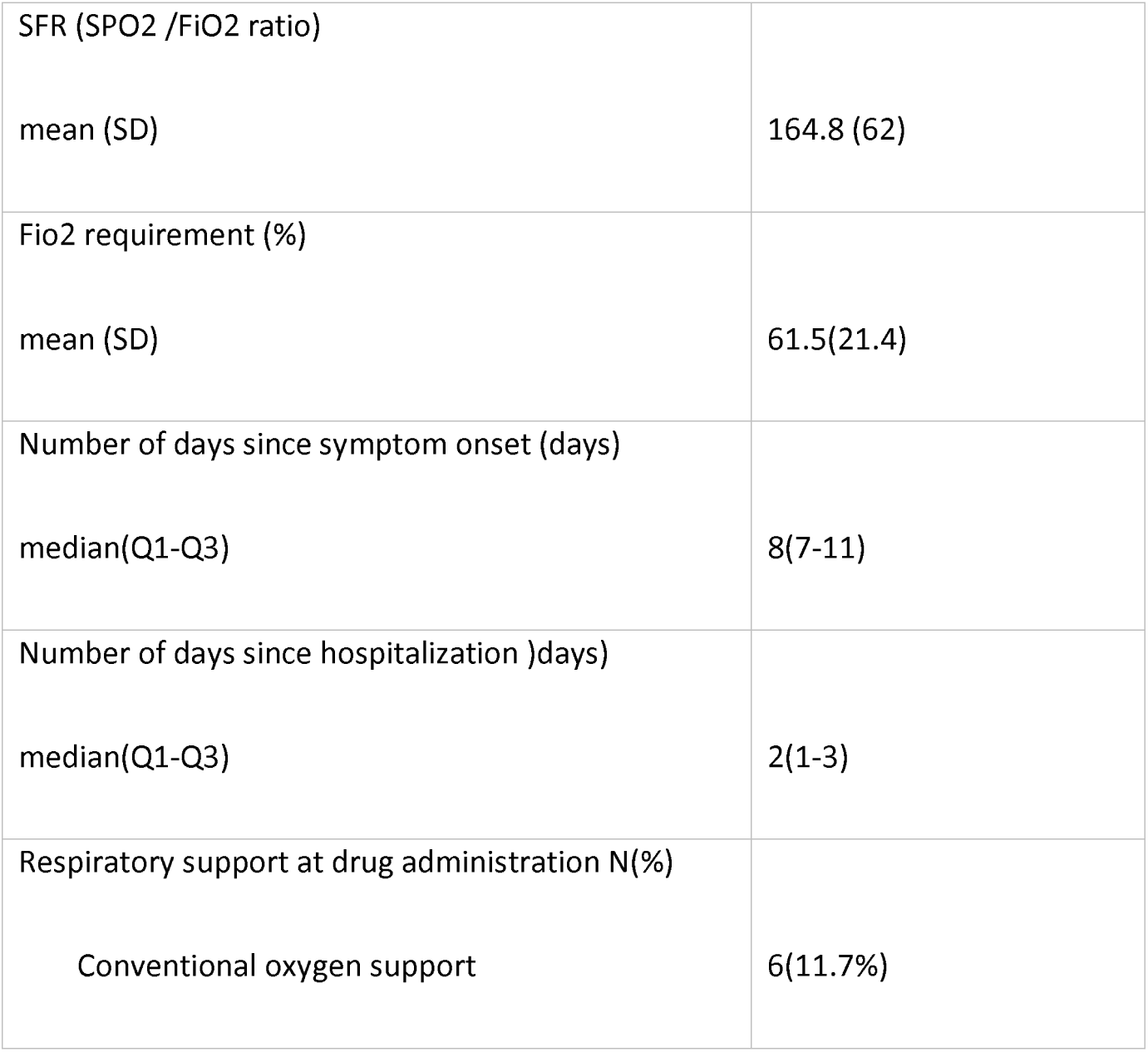
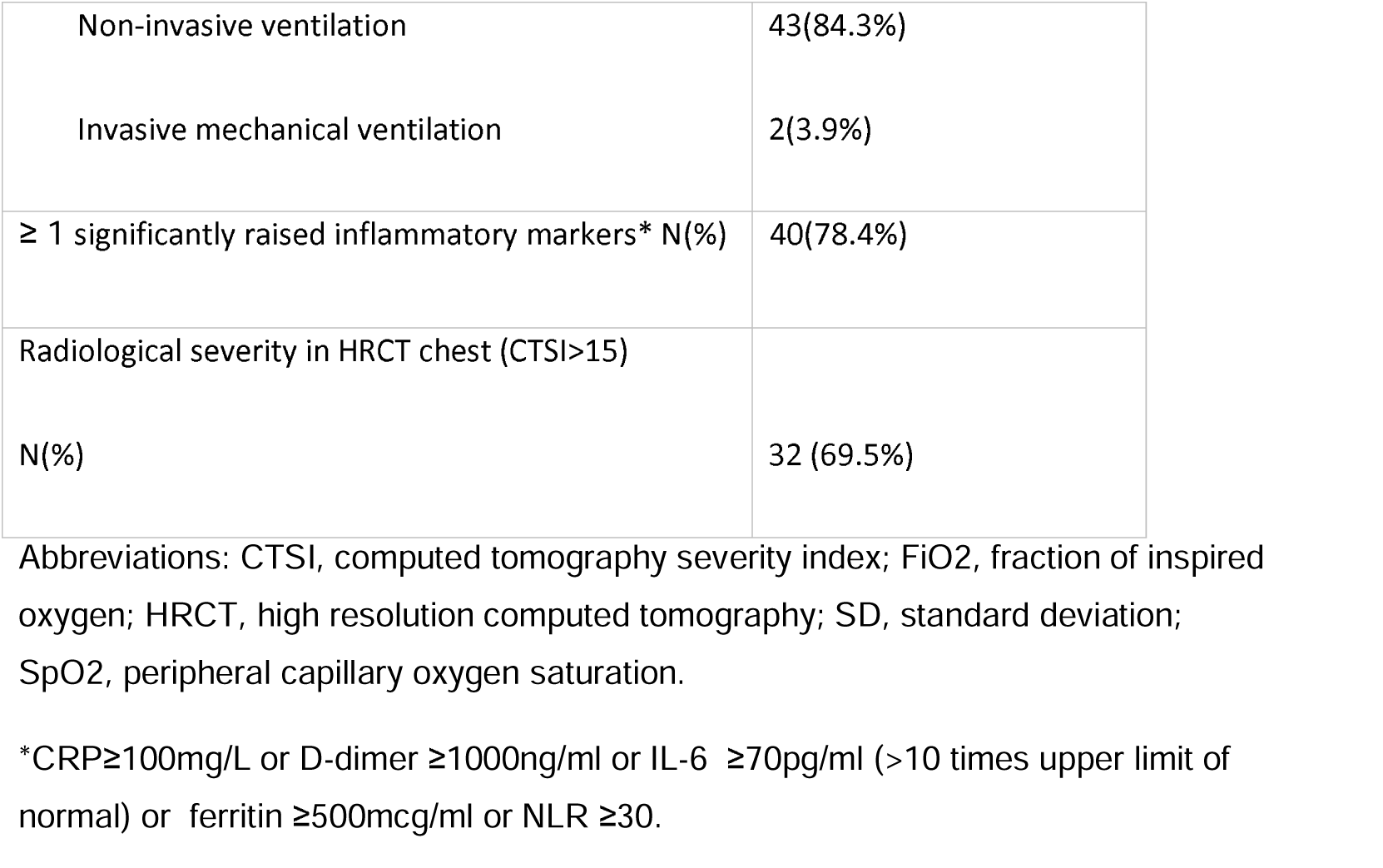
Other baseline clinical characteristics at tocilizumab administration

The median baseline values of various inflammatory markers on the day of tocilizumab administration and their trends are given in Table 5. Out of these only CRP showed a significant decrease on day 3 and day 7 after tocilizumab administration. However, CRP significantly decreased (p<0.001) in all patients irrespective of their outcome but the rest of the inflammatory markers showed a increasing trend (not significant) in those who had a poorer outcome (figure 1).

**Fig. 1.**
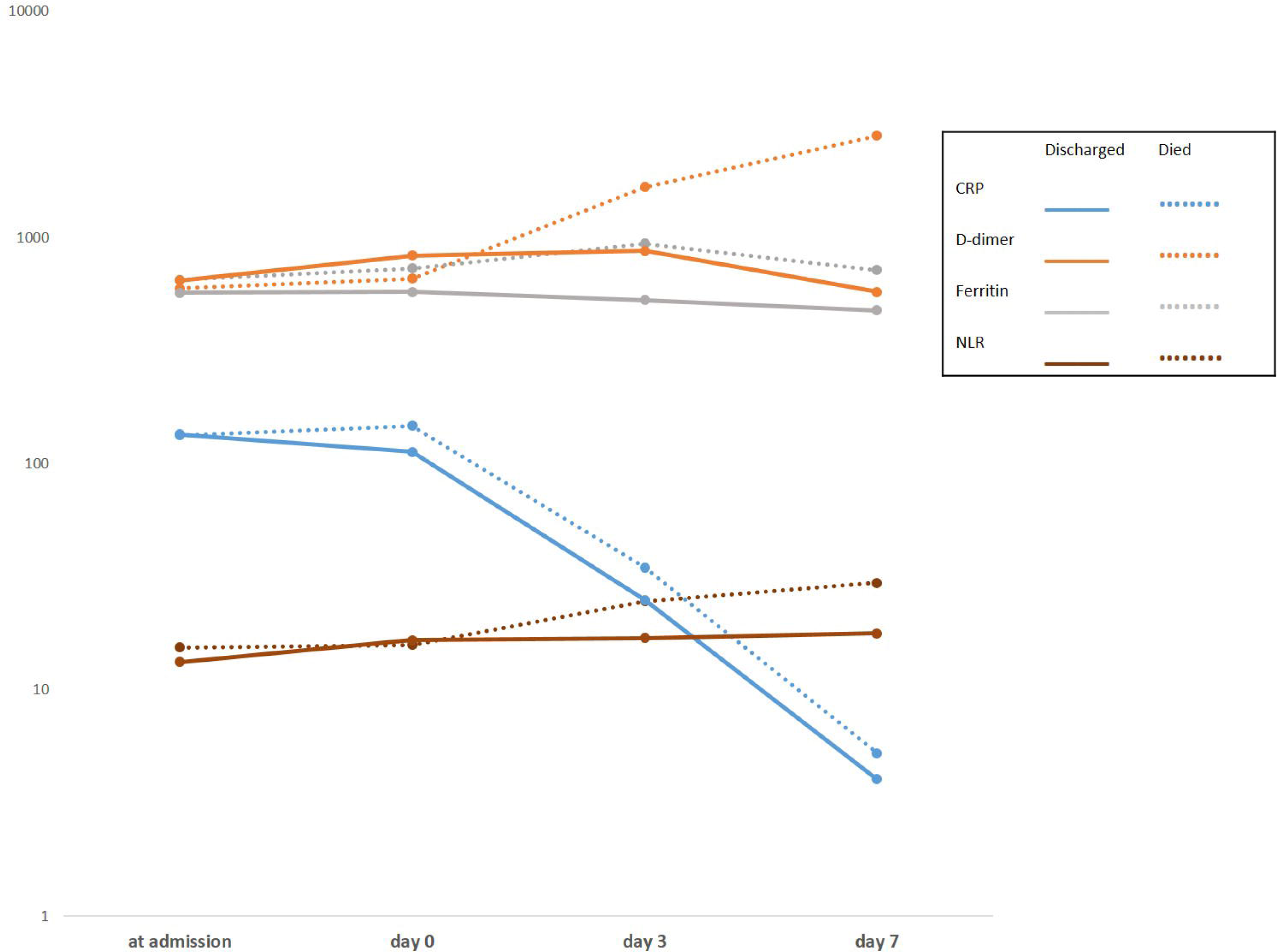

**Table 5.**
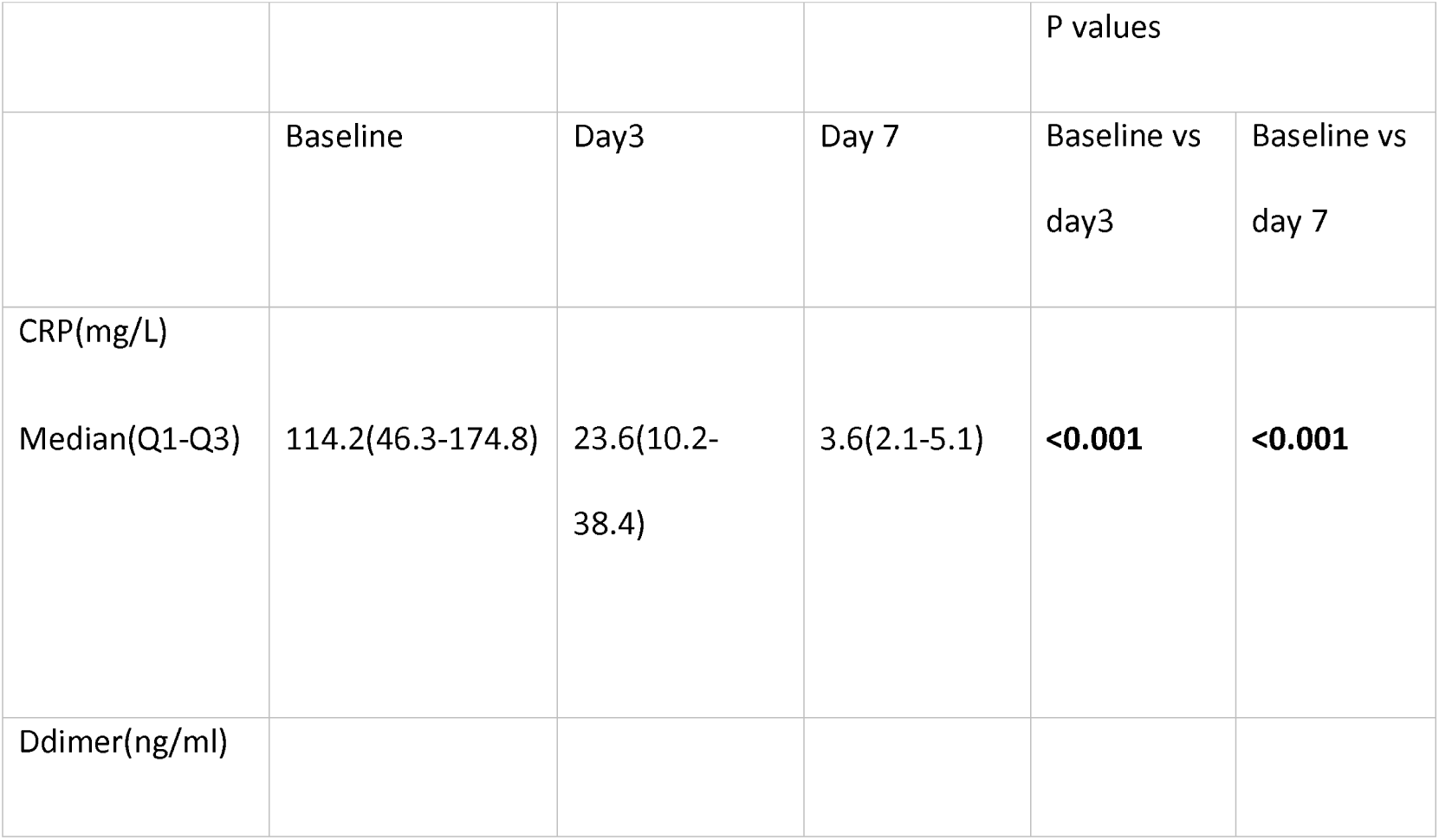
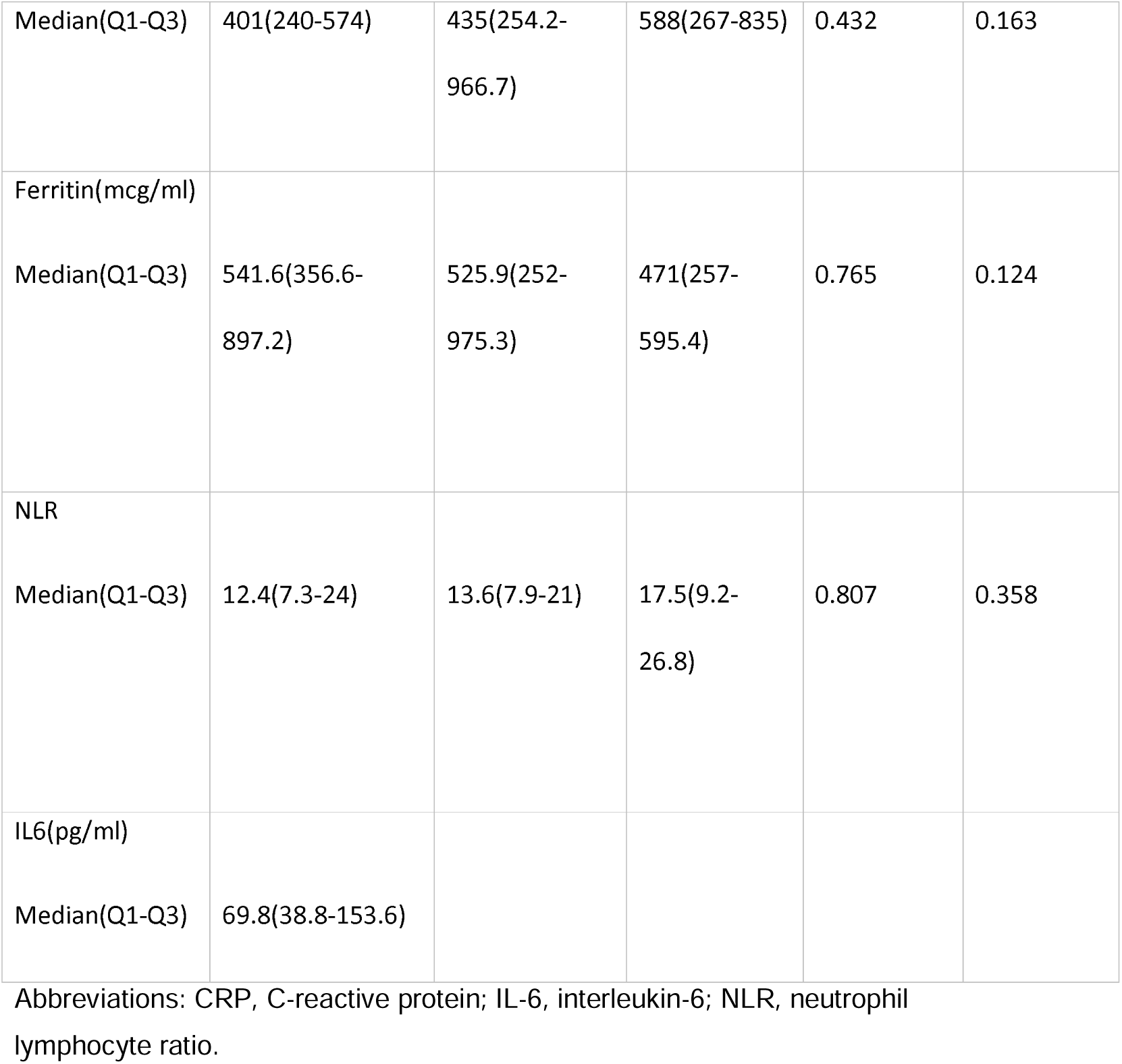
Trend in Inflammatory markers (median with interquartile ranges)

Following Tocilizumab administration, there was a significant decrease in oxygen requirement(p<0.001) and clinical progression (p-0.016) (by WHO clinical progression scale) by day 7(Table 6). Median time to oxygen independence from tocilizumab administration was 7.5 days (IQR,5-14). By day 28, 13/49(26.5%) patients who were on non-invasive oxygen support at the drug administration progressed to mechanical ventilation. Out of the intubated patients 5/15(33.3%) were successfully weaned of the ventilator. The all-cause mortality at day 28 was 10/51 (19.6%) (Table 7).

**Table 6.** O2 requirement and CPS (Clinical Progression Scale) score after tocilizumab administration

|  | Day 0 | Day 3 | Day 7 | Day 14 | Day28 |
| --- | --- | --- | --- | --- | --- |
| FiO2 (%) |  |  |  |  |  |
| mean(SD) | 61.5(21.4) | 57(24) | 43.6(25.9) | 32.8(22.3) | 25.4(13.2) |
| P values with respect to day 0 |  | **0.147** | **<0.001** | **<0.001** | **<0.001** |
| CPS score |  |  |  |  |  |
| Mean(SD) | 5.96(0.53) | 6(0.9) | 5.37(1.74) | 4.73(2.76) | 3.88(3.41) |
| P values with respect to day 0 |  | 0.569 | **0.016** | **0.032** | **0.01** |

Abbreviations: CPS, clinical progression scale; Fio2, fraction of inspired oxygen; SD, standard deviation.

**Table 7.** Primary outcome by day 28

|  |  |
| --- | --- |
| All-cause mortality N(%) | 10(19.6%) |
| Progression to mechanical ventilation N(%) | 13(26.5%)<sup>#</sup> |

<sup>#</sup>2 patients who were already on the ventilator while receiving the drug were excluded

More than 2/3rd (78.4%) of the patients received two doses(800mg) of tocilizumab. Overall 10/51(19.6%) patients had new onset of serious life threatening infections which included: bloodstream infections (4), Ventilator associated pneumonia (4), Hospital acquired pneumonia (3) and empyema (1). In 18/51 (35.3%) patients, antibiotics were escalated empirically at the discretion of the treating physician. Additionally, 5/51(9.8%) patients had thrombocytopenia, 3/51(5.9%) had pneumo-mediastenum or pneumothorax, 1/51(2%) patient had colonic perforation and 1/51(2%) patient had transaminitis (enzyme level >5 upper normal limit).

8/10 (80%) patients with ≥1 life threatening infections (as mentioned above) post tocilizumab administration and 10/15(66.7%) patients who were intubated during the ICU stay died at or before day 28 from the drug administration. Further on logistic regression after adjusting for age, comorbidities and baseline oxygen requirement, incidence of life threatening infections [aOR(adjusted odd ratio)-13.38,p-0.05) and intubated status(aOR-56.37,p-0.001) strongly predicted day 28 mortality after tocilizumab administration(Table 8)

**Table 8.**
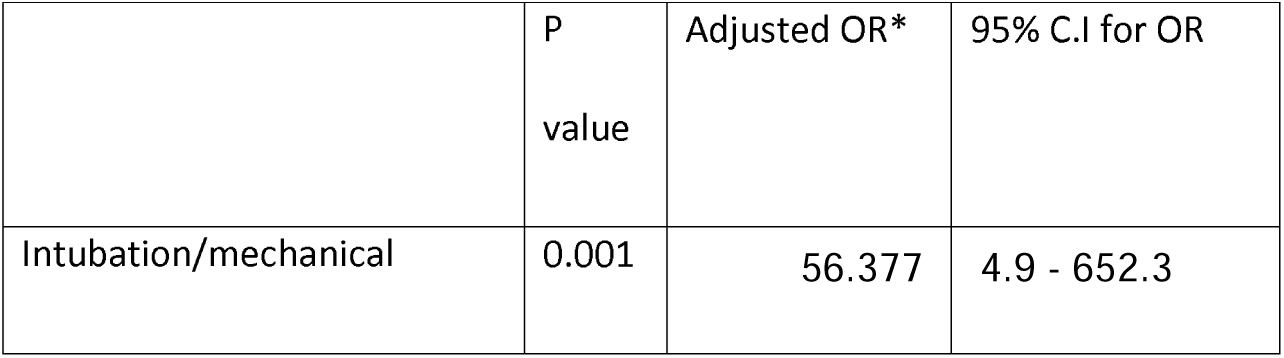
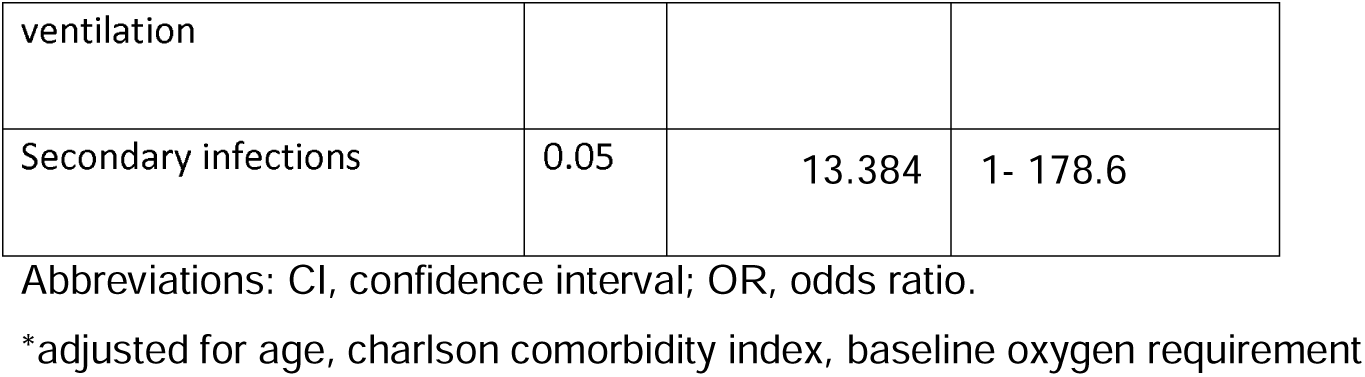
predictors of day 28 mortality at tocilizumab administration

## Discussion

Analysis of the above results indicates the usefulness of early and timely administration of tocilizumab in selected severe and critical COVID 19 patients who subsequently worsen or does not respond clinically with initial recommended doses of steroids. This approach can result in significant decrease in clinical progression and O2 requirement by the end of first week of treatment.

Most of the initial trials had predominately shown no benefit from IL-6 antagonists in COVID-19.^[12-14]^This may be due to the inclusion of less severely ill patients and exclusion of patients receiving any form of respiratory support. Moreover, only minority of the patients in these trials received tocilizumab in combination with corticosteroids. Recently published study by the REMAP-CAP investigators(n=865), included more critical patients, with 70.3% patients on noninvasive ventilation respiratory support and 29% on invasive mechanical ventilation with a median PaO2:FiO2 ratio(PFR) value of 116.5.^[15]^ The majority of patients (75.2%) in this study also received glucocorticoids at enrollment or within the following 48 hours. The in-hospital mortality in this study was 27% with IL6 receptor antagonist group vs 36% in the control group and they concluded that, the treatment with the IL6 receptor antagonists tocilizumab and sarilumab improved outcomes, including survival. The RECOVERY trial is the largest study till date(n-4116) on tocilizumab in COVID-19. This trial also included critical COVID-19 patients (40.9% on non-invasive and 13.6% on invasive respiratory support) and 82% of patients received steroids. This study showed an all cause mortality of 31% in tocilizumab group vs 35% in standard care group at day 28.^[16]^ Further a recent meta-analysis combining all the RCTs till date on tocilizumab on COVID-19 showed a significant benefit in critically ill subgroup of patients who are admitted to ICU (all cause mortality rate was 34.7% vs 39.6%).^[17]^ Even though a direct comparison of our data to these previous studies is not possible, the 28 day mortality rate in our patients (19.6%) shows a decreasing trend. This may be because we administered tocilizumab more selectively to ICU patients not responding to steroids rather than giving tocilizumab to all critically ill patients. This approach needs further validation with controlled trials. However, compared to the RECOVERY study, The progression to mechanical ventilation was much larger in our patients (26.5%), this may be because 84.9% of the patients in our group were already on non-invasive respiratory support on administration of tocilizumab as compared 41% in RECOVERY trial. We believe that our patients were more critical than the patients included in all the above studies due to the fact that steroid non responders were already having a poor clinical outcome. This is also reflected in the mean SpO2/FiO2(SFR) of 164.2(SD-62) [corresponding to moderate acute respiratory distress syndrome (ARDS)] at the time of administration of tocilizumab in our patients. The SFR is a noninvasive surrogate of PFR with good sensitivity and specificity.^[18]^ Moreover 29.4% of our patients were on mechanical ventilation on or before day 28, which predicted high mortality.

We obtained good results, even though tocilizumab in all our patients was administered only on the basis of clinical worsening. This may be due to the following reasons: Firstly, Inflammatory markers like CRP and IL-6 may be very sensitive, but their time to peak serum levels and half lives are not clear. Moreover, recent studies have shown that systemic levels of cytokines in COVID-19 may not be as high as seen with other causes of sepsis and ARDS.^[19]^ It may be that local inflammation, as evidenced by respiratory dysfunction, is a more useful indicator of which patients will benefit from IL-6 inhibition. Secondly, the CRP levels also decrease significantly after starting steroids,^[20, 21]^ which is also mirrored in our study, hence its level after initiating steroids may be unpredictable. Thirdly, as the clinical worsening occurs prior to radiological changes, we did not repeat chest radiology (if the previous imaging was recent(<24 hrs). Finally, the time lag in getting these reports, would mean a further delay in the drug administration.

Data from our patients shows that the trend in CRP is a poor prognostic marker for disease progression and outcome in those receiving tocilizumab and steroids. However, other inflammatory markers like D-dimer, ferritin and NLR can be valuable for the above purpose. This needs to be further confirmed in future trials.

As compared to previous studies, the incidence of life threatening secondary infection (19.6%) was high in our patient cohort. This was because combined use of steroids and tocilizumab make these patients vulnerable to severe secondary infection. This is a point of concern since 80% of these septic patients had poor outcome. Hence, a high number of patients (35.3%) needed empirical antibiotic escalation at the discretion of the treating clinician. This may be because, the classical clinical manifestations as well as the diagnostic accuracy of the laboratory chemical parameters of ongoing infections may be low in view of immunosuppression from steroids and tocilizumab.

Our study has some limitations. Firstly, the descriptive nature and absence of a comparator arm allows us to draw no meaningful conclusions about the population and is the major drawback of the study. Moreover, a comparator arm was difficult in the acute pandemic setting, in view of unavailability of other treatment regimens for the above clinical scenario and all the indicated patients had received the drug. Secondly, the number of patients observed were very less, thus generalization of the data is not possible. Finally, because of the short follow up period, we were not able to assess the long term safety and adverse effects of tocilizumab.

## Conclusion

To conclude, selective and timely administration of tocilizumab should be considered in severe and critical Covid 19 patients not responding to steroids. This can result in significant improvement in clinical progression and oxygen requirement. When given along with steroids, a high suspicion for secondary infections should be kept.

## Data Availability

All data produced in the present study are available upon reasonable request to the authors

## Acknowledgments and credits

none

